# An extended catalytic model to assess changes in risk for multiple reinfections with SARS-CoV-2

**DOI:** 10.1101/2023.09.27.23296231

**Authors:** Belinda Lombard, Cheryl Cohen, Anne von Gottberg, Jonathan Dushoff, Cari van Schalkwyk, Juliet R.C. Pulliam

## Abstract

**Background:** The SARS-CoV-2 pandemic has illustrated that monitoring trends in multiple infections can provide insight into the biological characteristics of new variants. Following several pandemic waves, many people have already been infected and reinfected by SARS-CoV-2 and therefore methods are needed to understand the risk of multiple reinfections.

**Objectives:** In this paper, we extended an existing catalytic model designed to detect increases in the risk of reinfection by SARS-CoV-2 to detect increases in the population-level risk of multiple reinfections.

**Methods:** The catalytic model assumes the risk of reinfection is proportional to observed infections and uses a Bayesian approach to fit model parameters to the number of *n*^*th*^ infections among individuals whose (*n* − 1)^*th*^ infection was observed at least 90 days before. Using a posterior draw from the fitted model parameters, a 95% projection interval of daily *n*^*th*^ infections is calculated under the assumption of a constant *n*^*th*^ infection hazard coefficient. An additional model parameter was introduced to consider the increased risk of reinfection detected during the Omicron wave. Validation was performed to assess the model’s ability to detect increases in the risk of third infections.

**Key Findings:** The model parameters converged when applying the model’s fitting and projection procedure to the number of observed third SARS-COV-2 infections in South Africa. No additional increase in the risk of third infection was detected after the increase detected during the Omicron wave. The validation of the third infections method showed that the model can successfully detect increases in the risk of third infections under different scenarios.

**Limitations:** Even though the extended model is intended to detect the risk of *n*^*th*^ infections, the method was only validated for detecting increases in the risk of third infections and not for four or more infections. The method is very sensitive to low numbers of *n*^*th*^ infections, so it might not be usable in settings with small epidemics, low coverage of testing or early in an outbreak.

**Conclusions:** The catalytic model to detect increases in the risk of reinfections was successfully extended to detect increases in the risk of *n*^*th*^ infections and could contribute to future detection of increases in the risk of *n*^*th*^ infections by SARS-CoV-2 or other similar pathogens.

## Introduction

In March 2020, a global pandemic of Coronavirus-Disease-19 (COVID-19), caused by severe acute respiratory syndrome coronavirus 2 (SARS-CoV-2), was declared by the World Health Organisation (WHO) (1). During the COVID-19 pandemic, numerous mathematical models were created to estimate the SARS-CoV-2 reproduction number, investigate the influence of implemented public health interventions on the transmission dynamics, and assess the global spread of the disease (2). It is becoming increasingly important to study the risk of multiple reinfections, especially in the context of waning immunity or the emergence of different viral variants. Recovered individuals could be a source of spread of SARS-CoV-2 and knowing the risk of getting reinfected by SARS-CoV-2 is therefore important.

Prior research, such as the study by Wangari *et al*. examined reinfection transmission mechanisms using a compartmental model (3). Another model was developed to validate a test-negative study design to rapidly and rigorously estimate the protection conferred by prior infection (4); this design was applied to data in Qatar and found that protection against reinfection was higher when the primary infection was caused by the Alpha variant compared to the Beta variant (4).

Another model used to study reinfection dynamics was a catalytic model by Pulliam *et al*. (5). In this study, the model parameters were fitted to reported reinfections of SARS-CoV-2 up to a specific date. Reinfection numbers were then projected under the assumption of constant reinfection risk and compared to observed data during a projection period to determine whether this assumption had been violated. The model assumed that the reinfection hazard was proportional to the seven-day moving average of detected cases, with a constant hazard coefficient (5). The 95% credible interval of projected reinfections was compared to the observed reinfections during the projection period to assess whether a change in the reinfection hazard coefficient had occurred (5). The study identified a deviation from the projections during the Omicron wave in November 2021, providing the first evidence of higher risk of reinfection by the Omicron variant than previous variants (5).

All these studies focused on monitoring the risk of second infection. During the Omicron wave in South Africa there was a noticeable increase in the number of third infections. Since multiple reinfections (three or more infections) are becoming more prevalent, our study extends the model developed in Pulliam *et al*. to detect increases in the risk of multiple reinfections in South Africa. The original model findings, complemented by the findings from the extended model have been applied to South African data and published in the National Institute for Communicable Diseases (NICD) monthly report on SARS-CoV-2 Reinfection Trends in South Africa (6).

By providing insights into the risk of multiple reinfections, our study contributes vital information to the SARS-CoV-2 literature body to guide public health decisions, specifically relating to interventions used to prevent spread, such as vaccination (7). The extended model could be used to identify a new emerging immune-escaping variant of SARS-CoV-2, which would consequently guide policy surrounding an outbreak.

## Methodology

### Data source

The dataset used in this study is a time-series of the daily counts of primary infections, second infections, third infections and fourth infections of SARS-CoV-2 in South Africa from 4 March 2020 to 29 November 2022. This dataset, as detailed in Pulliam *et al*. (5) is accessible on Zenodo (DOI: 10.5281/zenodo.7426515). If the model is used for the risk of an *n*^*th*^ infection (where *n* can be 3 or more), the column with the number of observed *n*^*th*^ infections must be included in the dataset.

The observed infections in the dataset (5) were obtained from a national dataset containing all positive tests in South Africa, detected with either Polymerase Chain Reaction (PCR) or rapid antigen. Reporting of positive tests by laboratories was mandatory, although antigen detection tests are known to have been underreported. In the dataset, deterministic and probabilistic linkage methods were used to identify repeated tests of the same person. To identify suspected reinfections (where reinfections can be second, third, or fourth infections), positive tests of an individual that were at least 90 days after the most recent positive test from the previously observed infection were identified. The specimen receipt date was used as the date of reference in the analysis. In this study, we focus on the number of third infections (*n* =3).

### The extended model

In the adapted model, the number of *n*^*th*^ infections expected on day *x* is calculated from the number of (*n* − 1)^*th*^ infections reported at least 90 days prior to day *x* and have not led to a detected *n*^*th*^ infection. As in Pulliam *et al*., a reinfection is defined as an individual having two positive tests for SARS-CoV-2 at least 90 days apart. This delay is introduced to distinguish reinfection from prolonged viral shedding, the latter being where an individual actively releases the virus (8,9) and may lead to a positive test even though they are no longer symptomatic or infectious (8).

Building on the original catalytic model, the equation to calculate the probability of an *n*^*th*^ infection by day *x*, given a previous (*n* − 1)^*th*^ reported infection on day *t* is described as

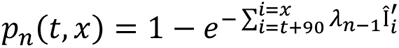

where 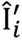 is the 7-day moving average of total infections reported on day *i, n* is the number of infections for which the risk is being studied and *λ*_*n*−1_ is *n*^*th*^ infection hazard coefficient being fitted to the data. The total reported infections on day *i* can be described as:

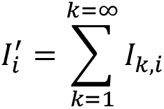

where *I*_*k,i*_ is the number of *k*^*th*^ infections reported on day *i*.

The expected number of *n*^*th*^ infections, *Y*_*n,x*_, reported by day *x* can therefore calculated as:

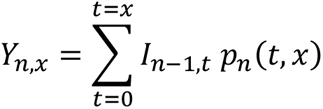

The expected number of *n*^*th*^ infections on day *x* can then be calculated as:

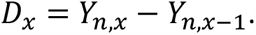

We used the extended model to assess third infection risk in South Africa from March 2020 to November 2022 and then subjected it to simulation-based validation to evaluate the performance of the method under a broad range of scenarios. The model was implemented in the R Statistical Programming Language [version 4.3.1 (2023-06-16)] (10). The code for the extended model is available on Github at https://github.com/SACEMA/reinfectionsBelinda.

### Fitting the extended model to South African data on third infections (*n* = 3)

Two parameters, the hazard coefficient for *n*^*th*^ infections, *λ*_*n*−1_, and the negative binomial dispersion parameter, *κ*_*n*−1_ were fitted to the data using Monte Carlo Markov Chains (MCMC) with 10,000 iterations and four chains, with 1,500 iterations discarded as burn-in for each chain. Convergence of the parameters was measured using Gelman-Rubin diagnostics with the ‘gelman.diag’ function from the *coda* package in R (11). Gelman-Rubin compares the within-chain variance and the between-chain variance to evaluate the Monte Carlo Markov Chains, as this gives an indication of whether the initial value has been “forgotten”. A value of less than 1.1 indicates a low difference between the variances and, therefore, convergence (12,13).

The projected *n*^*th*^ infections were calculated from a joint posterior distribution from the chains that were fitted during the MCMC procedure, so that 2,000 samples were drawn over the four chains equidistance apart (after discarding burn-in). For each model parameter combination from the posterior distribution, 100 stochastic simulations were run to calculate the number of expected third infections for each day for that parameter combination. From the realisations, two 95% projection intervals are calculated: the middle 95% of the expected daily *n*^*th*^ infections, and the middle 95% of the 7-day moving average of expected *n*^*th*^ infections.

### Fitting with an additional parameter to third infections data (*n* = 3)

An increase in the risk of a second infection was detected by Pulliam *et al*. during the Omicron wave. Model parameters did not converge when considering the risk of third infections (*n* =3) due to low observation of third infections before the Omicron wave. To overcome this issue, the fitting period was extended beyond October 2021 and an additional parameter was introduced to account for the increased risk of a second infection with this variant (4).

The fitting period for 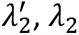 and *κ*_2_ to detect increases in the risk of third infections was extended to include the first Omicron wave, up to 31 January 2022.

The probability of having a third reported infection by day *x*, given that the person had a positive test for a second infection on day *t*, can then be given by:

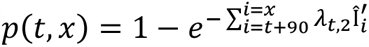

where

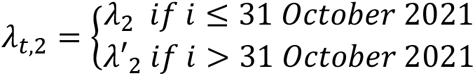

### Model validation

In (14), we conducted simulation-based validation to assess the performance of the original catalytic model when introducing changes in the risk of second infections under different scenarios.

We concluded that the model is robust to several important aspects of the observation process that are not directly accounted for in the model. Here, we assessed the model by performing sensitivity analyses on the model’s suitability for assessing third infections under different increases in the risk of second and third infections.

To validate the *n*^*th*^ infection method proposed in this study, we considered a simulated dataset of primary infections that represents perfect observation (adapted from the data as in (14)), on which we added fixed primary infection, second infection, and third infection observation probabilities (0.2, 0.5 and 0.35 respectively). The lower third infection observation probability was considered to account for pandemic fatigue (15). We tested the performance of the model on simulated data in different data-generation scenarios by varying the difference in reinfection risk (both the second infection and third infection risk) between a pre-Omicron-like period and from when an Omicron-like wave is introduced. This approach determined whether the model could accurately fit the model parameters for third infections (*λ*_2_, 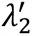 and *κ*_2_) and whether the model gives a false positive and detects increases in the risk of third infection where there is not such an increase.

From the simulated primary infections, we generated a time-series for the number of observed primary infections by drawing a binomial random variable based on the observation probability, so that

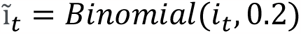

where Ĩ_*t*_ is the observed primary infections on day *t* and *i*_*t*_ is the number of underlying primary infections on day *t*.

From the number of observed primary infections, the number of underlying second infections per day was calculated as:

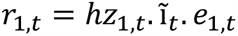

where *e*_1_,_*t*_ is the underlying number of people eligible for second infection and calculated as

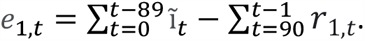

The modified second infection hazard coefficient, *hz*_1,*t*_, is calculated as:

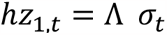

where Λ is the second infection hazard coefficient and *σ*_*t*_ is a multiplier on the coefficient (scale parameter) to represent the increase in second and third infection risk, and defined as

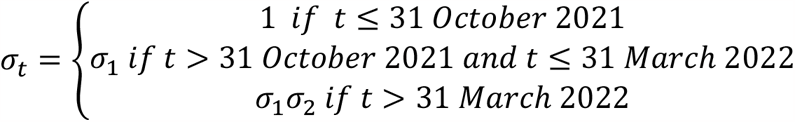

We used *σ*_1_ to represent the first increase in second and third infection risk associated with the Omicron wave and introduced an additional parameter *σ*_2_ to evaluate whether the method can detect additional simulated increases in the risk of a third infection. Table 1 shows how *σ*_1_ and *σ*_2_ were varied. In the second scenario, we fixed *σ*_1_ at 2.8, which was the median of the ratio of the posteriors of 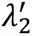 and *λ*_2_ obtained from fitting the parameters to the third infection data in South Africa.

**Table 1.** The values of σ_1_ and σ_2_ considered in the data simulation process for the data that was used in the simulation-based validation.

| Scenario | Values of $\sigma_1$ | Values of $\sigma_2$ |
| --- | --- | --- |
| 1 | {1, 1.2, ..., 3} | {1} |
| 2 | {2.8} | {1, 1.2, 1.5, 2} |

We fixed Λ as the median of the fitted second infection hazard coefficient (*λ*) estimated in Pulliam *et al* (as the median of the posterior distribution of *λ* obtained).

The number of observed second infections on day *t* was calculated as

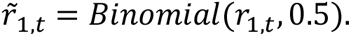

This was further extended to calculate the number of underlying third infections on day *t, r*_2,*t*_ as

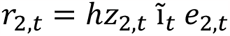

where *hz*_2,*t*_ is the third infection hazard coefficient and *e*_2,*t*_ is the number of people eligible for a third infection on day *t*, calculated as

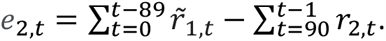

We calculated *hz*_2,*t*_ in a similar way as *hz*_1,*t*_:

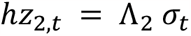

where Λ_2_ is the third infection hazard coefficient. We fixed Λ_2_ at approximately 6.6 · 10^−8^, which is obtained from the median of the posterior distribution of the third infection hazard coefficient 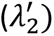 that were fitted with the observed third infections in South Africa during the Omicron wave.

The number of observed third infections was calculated as

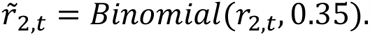

In the data generation process, we varied *σ*_*t*_ according to the scenarios described in Table 1. For each of these values of *σ*_1_ and *σ*_2_, a time-series of second and third infections was generated using the process described and applied to the model fitting (fitting *λ*_2_, 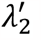 and *κ*_2_) and projection process. The process was repeated 20 times with different seeds. We measured the Gelman-Rubin convergence diagnostics for *κ*_2_, *λ*_2_ and 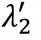. We also measured the proportion of observed third infections that were above the 95% 7-day moving average during the projection interval, as well as the timing of the first cluster of five consecutive days of observed third infections that fell above the projection interval, denoted as *D*_*first*_, which can be used to detect increases in the risk of third infections.

In the case of 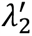 for third infections when *σ*_2_ = 1, we expect that around 2.5% of the observed third infections fall above the 95% projection interval, and we expect that a cluster of five consecutive days above the projection interval will not exist, since *σ*_2_ = 1 indicates that we did not introduce a further increase in the risk of third infections in the simulated data after 31 October 2021. The existence of such a cluster may indicate a false positive detection of an increase in the risk of third infections. We therefore measured the specificity of the model for each value of the scale parameter *σ*_1_ (with *σ*_2_ = 1) as

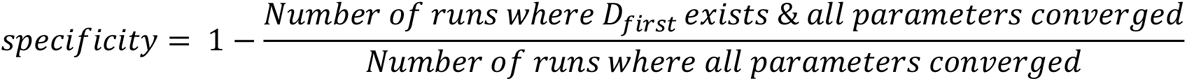

## Results

### Data used in the third infections fitting procedure

In Figure 1, the number of primary, second, and third infections reported in South Africa from 4 March 2020 to 29 November 2022 is depicted. Figure 1A shows the number of observed primary infections, Figure 1B shows the number of people eligible for a second infection, Figure 1C shows the number of observed second infections, Figure 1D shows the number of people eligible for a third infection, and Figure 1E shows the number of observed third infections.

**Figure 1.**
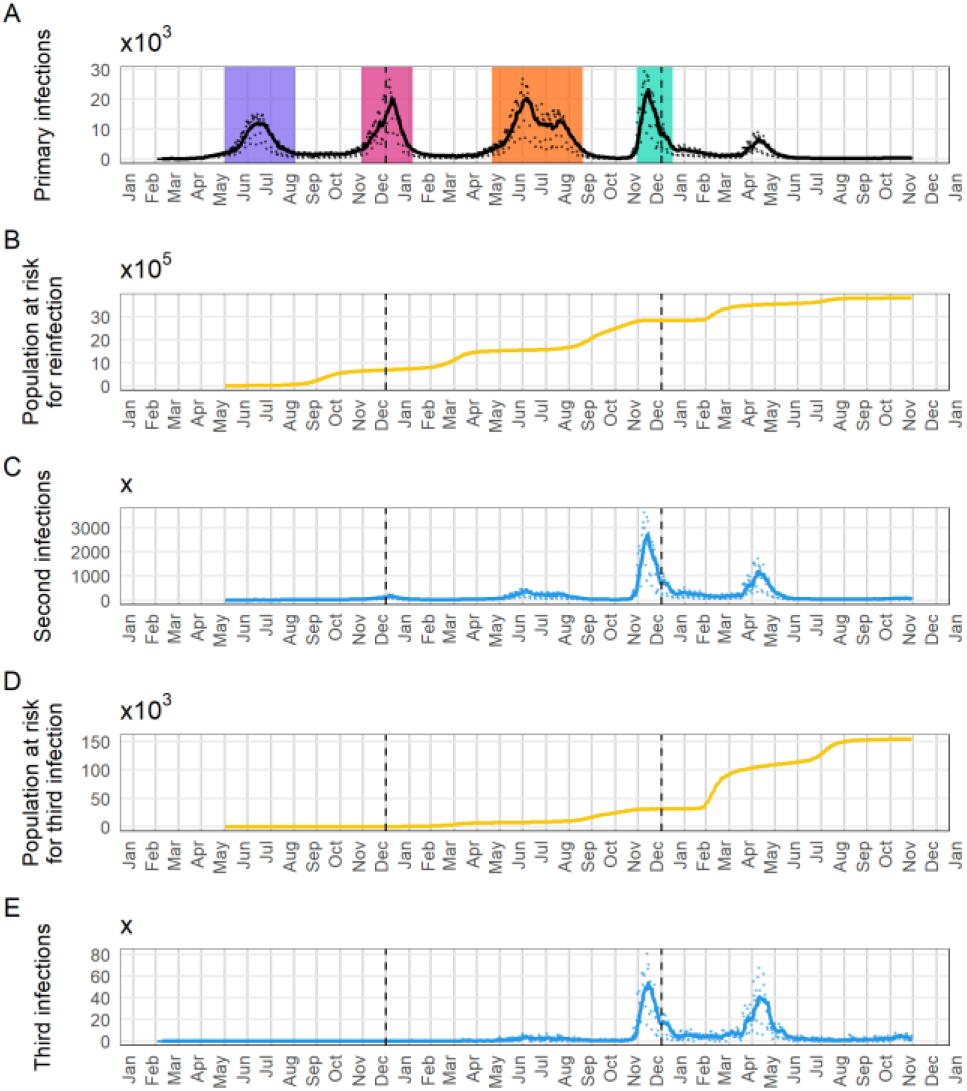
Infections reported to the NICD in South Africa and the number of people eligible for second and third infections (calculated from the reported infection data), from 4 March 2020 to 29 November 2022. Figure 1A shows the number of observed primary infections, Figure 1B shows the number of people eligible for a second infection, Figure 1C shows the number of observed second infections, Figure 1D shows the number of people eligible for a third infection, and Figure 1E shows the number of observed third infections. Source: https://zenodo.org/record/7426515.

### Model fitting

With only the third infection hazard coefficient, *λ*_2_, and the negative binomial dispersion parameter, *κ*_2_, fitted to third infections up to October 2021, *λ*_2_ converged well (convergence diagnostic of ∼1.03), while *κ*_2_ did not converge due to low numbers of third infections (convergence diagnostic of ∼1.3) (Figure S1**Error! Reference source not found**.).

The fitting period was then extended through the Omicron period up to 31 January 2022, and the second third infection hazard coefficient, 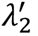, was introduced. When adding this third parameter to the fitting procedure, *λ*_2_, 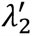 and *κ*_2_ converged well, with the Gelman-Rubin diagnostic values falling below 1.1. The convergence diagnostic for 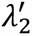 was slightly higher (around 1.05) than for *λ*_2_ and *κ*_2_ (approximately 1.01 and 1.005 respectively, Figure 2).

**Figure 2.**
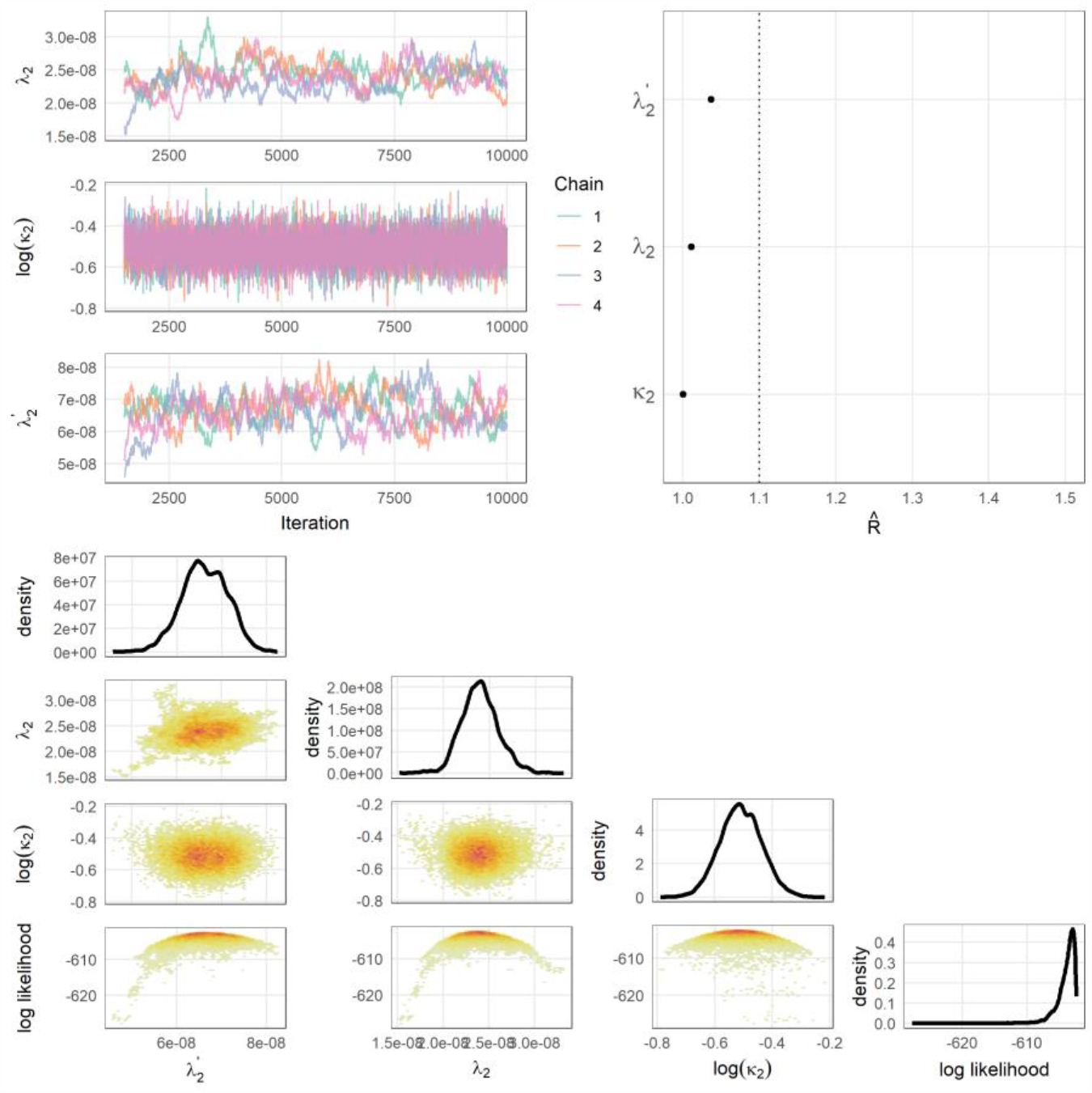
Convergence diagnostics plot when fitting λ_2_, 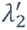 and κ_2_ to the data in South Africa. The top left panels show the trace plots for each parameter. On the right is the Gelman-Rubin convergence diagnostics. The plots at the bottom are density plots of the fitted parameters.

### Model prediction

Figure 3 and Figure S2 shows the 95% projection interval of expected third infections (both the 7-day moving average and the daily third infections) and the observed third infections when the model was fitted to South African data and used to project third infections. In Figure S2, the data were fitted with only *λ*_2_ and *κ*_2_, and the 7-day moving average of observed third infections (red solid line) did not stay in the 95% projection interval for most of the projection interval (the red band). The negative binomial dispersion parameter (*κ*_2_) did not converge in this scenario. In Figure 3, the additional third infection hazard coefficient parameter was also fitted 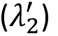, and the fitting period was extended to include the first Omicron wave. In this case, the observed third infections mostly stayed within the 95% projection interval. From May to November 2022, the number of observed third infections (red solid line) reaches the lower edge of the band of the 95% 7-day moving average projection interval of third infections (red band), showing a potential decreased risk of third infections, or a lower observation probability due to pandemic fatigue.

**Figure 3.**
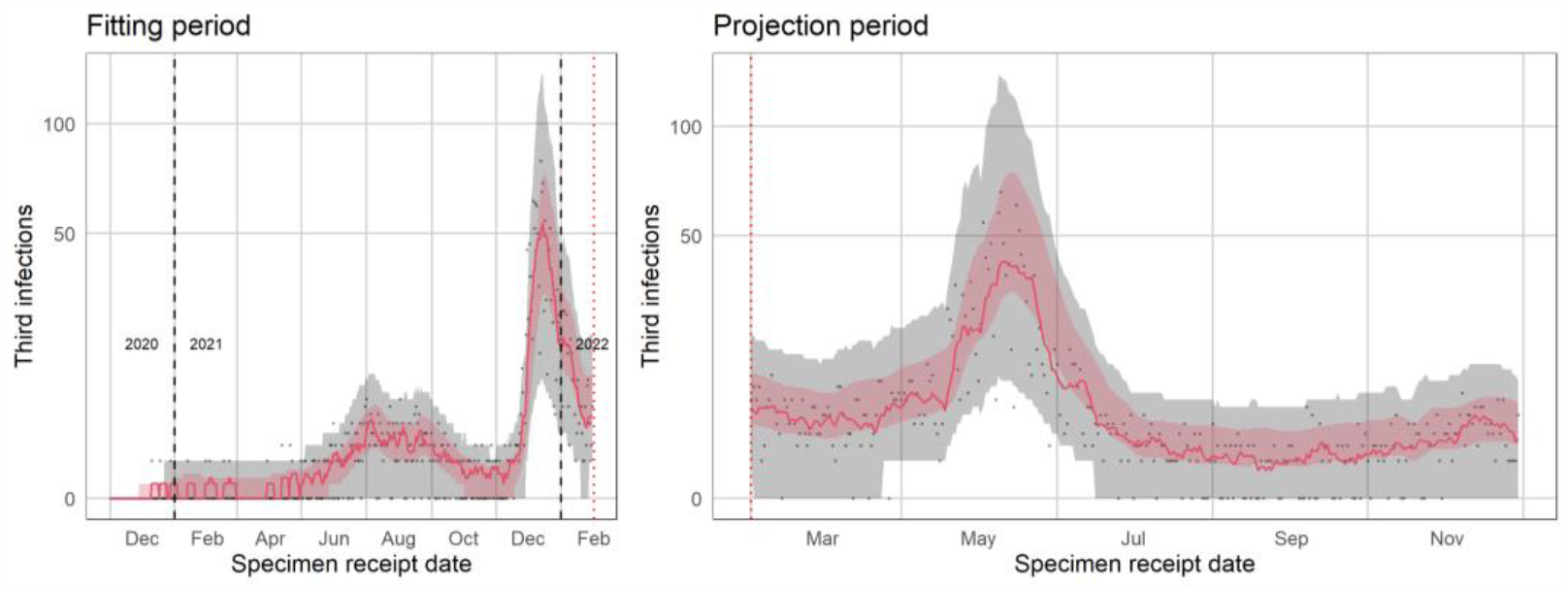
Simulation Plot published in the NICD reinfections report with the added third infection hazard coefficient to represent the Omicron wave 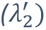 parameter and data fit through 31 January 2022. (https://www.nicd.ac.za/wp-content/uploads/2022/12/SARSCoV2-Reinfection-Trends-in-South-Africa_2022-12-07.pdf). The left side represents the fitting period, and the right-hand side the projection period. The red band is the 95% projection interval for the 7-day moving average of simulated third infections for that day and the grey band is for the daily simulated third infections. The red solid line is the 7-day moving average of observed third infections and the grey dots are the daily values of observed third infections.

### Model validation

After running the model fitting and model projection 20 times for each value of *σ*_1_ and fixing *σ*_2_ = 1, the negative binomial dispersion parameter (*κ*_2_) mostly converged when *σ*_1_ > 1.6. The proportion of runs where *κ*_2_ converged increased as *σ*_1_ increased, due to increased numbers of third infections. For more than 0.75 of the runs for each value of *σ*_1_ the third infection hazard coefficient before the first Omicron wave (*λ*_2_) converged, whereas the third infection hazard coefficient for after the first Omicron wave 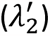 converged in all the runs (Figure S3).

The specificity (proportion of runs where an increase in the risk of third infection was not detected when there is no increase in third infection risk in the generated data, *σ*_2_ = 1) was above 0.75 for all values of *σ*_1_ and 1 when *σ*_1_ < 1.5 (Table S1).

After excluding the results from runs with nonconvergence, the proportion of observed third infections above the projection interval remained below 2.5%, except one run where *σ*_1_ =3 which resulted in 5% of third infections above the projection interval.

When fixing *σ*_1_ = 2.8 and varying *σ*_2_ with values of 1, 1.2, 1.5 and 2, the median of *D*_*first*_ from the runs where all the parameters (*κ*_2_, *λ*_2_ and 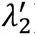) converged decreased from 26 days to 7 days as *σ*_2_ increased from *σ*_2_ = 1.2 to *σ*_2_ = 2 (Figure S4), and *D*_*first*_ did not exist in most cases where *σ*_2_ = 1 (specificity was 0.89). The proportion of points above the projection interval was 0.01 when *σ*_2_ = 1 and gradually increased to 0.45 when *σ*_2_ = 2 (Figure S4).

## Discussion

In this study, the method used to detect changes in the risk of reinfection was successfully extended to detect the risk of multiple reinfections. The output of the extended method for third infections was used by NICD in their monthly report to detect further changes in the risk of third infections (6) and will contribute to future monitoring of reinfection risk where there are concerns about potential emerging SARS-CoV-2 variants with immune escape and where multiple reinfections with SARS-CoV-2 are relevant. With the extended method, we have demonstrated that we would have detected increases in the reinfection risk during the fifth wave if such an increase existed.

We performed a simulation-based validation of the method, where simulated data on third infections with SARS-CoV-2 were fitted and projected. The model is robust to changes in the risk of third infections when we fitted an additional parameter that represents the second and third infection hazard coefficient during waves where the reinfection risk is higher. When the increase in the second and third risk in the simulated data used for the validation was low, the negative binomial dispersion parameter did not converge in some runs. This is due to an insufficient number of simulated third infections to properly inform the parameter, whereas with the higher increase in the risk of second and third infection, more data were generated to properly inform the dispersion parameter. The specificity, which assesses the method’s ability to avoid false positive detections of third infection risk increases during the projection period, was generally high for most scale values representing the initial rise in third infection risk (first Omicron wave). This suggests that the model effectively distinguished increases in the risk of reinfections from random fluctuations or noise in the data. When introducing an additional increase in the third infection risk after the additional hazard coefficient parameter is introduced), the method detects the simulated increase in the risk of third infection even for the smallest increase we investigated (*σ*_2_ = 1.2). The proportion of points above the projection interval after the introduction of the additional increase in third infection risk was only 45% when the increase was 100% (*σ*_2_ = 2), which could be due to the low number of observed third infections after the fifth wave. The low numbers of observed third infections are likely due to pandemic fatigue.

Validation has been performed when considering third infections, however when the model is used to study the risk of more than three infections, it is advised to conduct further validation. To ensure accuracy when using the model in future predictions about risk of reinfection, it is important to incorporate prior knowledge and additional parameters, such as introducing a third lambda parameter to account for changing reinfection risks, if necessary.

A limitation of the method is its sensitivity to low counts of observed reinfections, as sufficient reinfection is required for the model parameter convergence. Pandemic fatigue, which leads to fewer testing and consequently lower numbers of observed reinfections (reinfections beyond the second infection), impacts the method’s applicability in a real-life situation. With low numbers of observed multiple reinfections, the model is less likely to detect increases in the risks of multiple reinfections.

## Conclusion

The catalytic model used to detect increases in the risk of reinfection was successfully generalised to detect increases in the risk of *n*^*th*^ infections. The method was applied to the observed third infections in South Africa to detect increases in the risk of third infection, and simulation-based validation showed its robustness in detecting increases in the risk of third infections of different magnitudes. The extended method could contribute to future detection of increases in the risk of *n*^*th*^ infections by SARS-CoV-2 or other similar pathogens.

## Data Availability

All data produced and used are available online a
https://github.com/SACEMA/reinfectionsBelinda and https://zenodo.org/record/7426515

https://zenodo.org/record/7426515

https://zenodo.org/record/8354838

## Acknowledgements

J.R.C.P. and C.v.S. are supported by the South African Department of Science and Innovation and the National Research Foundation. Any opinion, finding, and conclusion or recommendation expressed in this material is that of the authors, and the NRF does not accept any liability in this regard. This work was also supported by the Wellcome Trust (grant no. 221003/Z/20/Z) in collaboration with the Foreign, Commonwealth and Development Office, United Kingdom. Writing assistance was provided by Yuri Munsamy, PhD of SACEMA, South Africa. This assistance was funded by Stellenbosch University. The authors gratefully acknowledge the Centre for High Performance Computing (CHPC), South Africa, for providing computational resources to this research project.

This work has benefited from input during the Software for the Applied Mathematical Sciences (SEAMS) workshop which is part of the International Clinics on Infectious Disease Dynamics and Data (ICI3D) program. We specifically thank C. A. B. Pearson, T.J. Hladish, A. Stoltzfus for helpful discussions during the development of this work.

We are grateful to Nevashan Govender, Michelle J. Groome, Koleka Mlisana and Harry Moultrie for their ongoing support of work on SARS-CoV-2 reinfections in South Africa.

## Supplementary material

**Figure S1.**
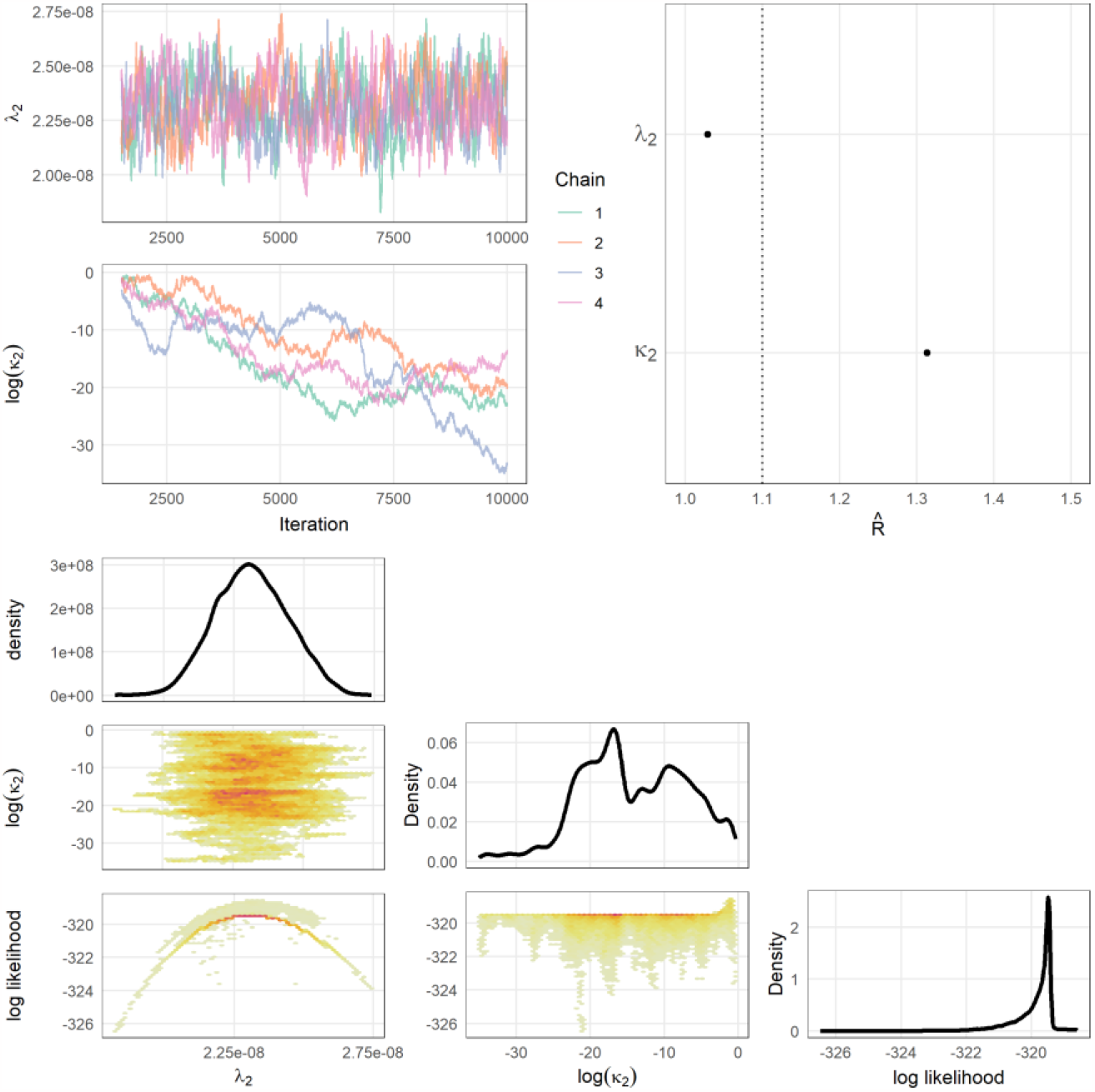
Convergence diagnostics with λ_2_ and κ_2_ fitted to the data. The top left panels show the trace plots for each parameter. On the top right is the Gelman-Rubin convergence diagnostics. The plots at the bottom are density plots of the fitted parameters. The fitting period is up until 31 October 2021 (before the Omicron period).

**Figure S2.**
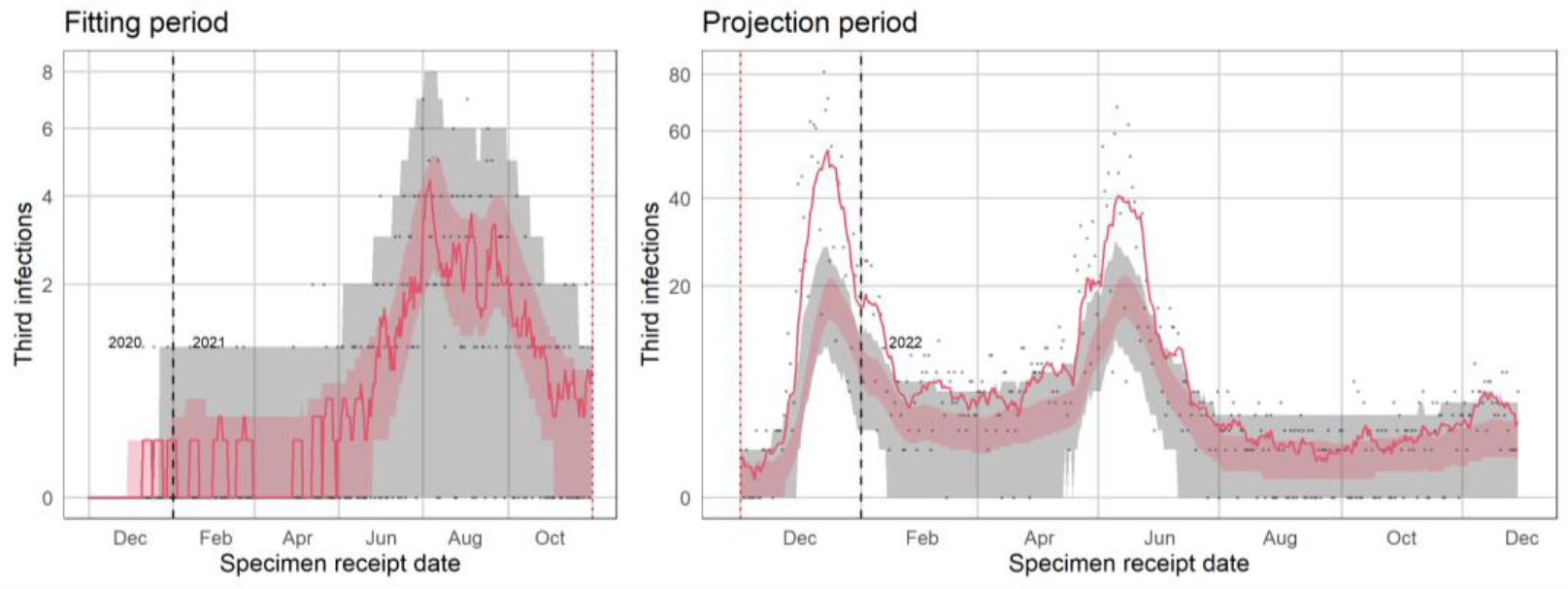
Simulation Plot for when only λ_2_ and κ_2_ were fitted up until 31 October 2021 and projected through the Omicron wave. The left side represents the fitting period, and the right-hand side the projection period. The red band is the 95% projection interval for the 7-day moving average of simulated third infections for that day and the grey band is for the daily simulated third infections. The red solid line is the 7-day moving average of observed third infections and the grey dots are the daily values of observed third infections.

**Figure S3.**
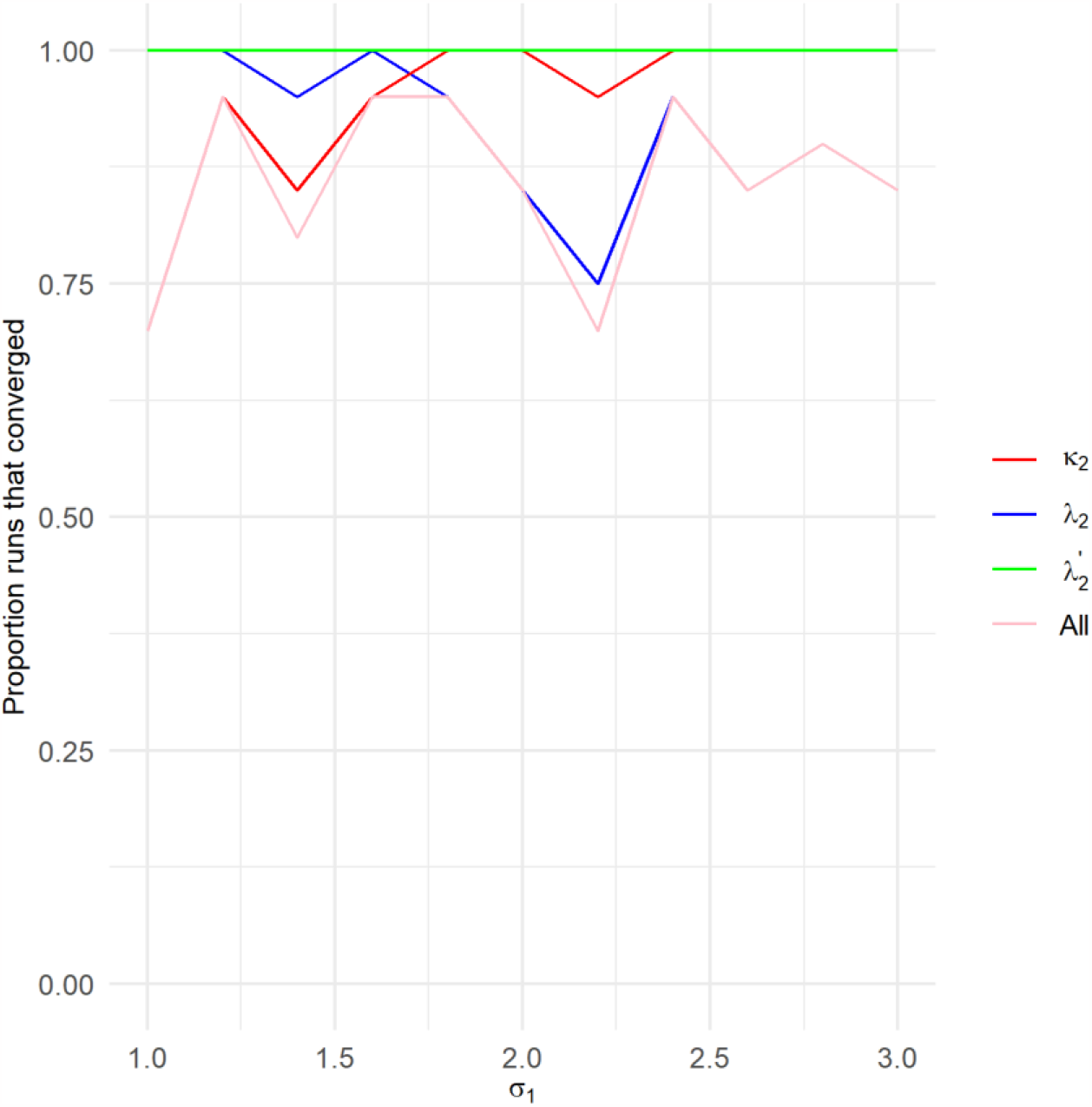
The proportion of the 20 runs in which κ_2_, λ_2_ and 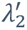 converged respectively. In this plot, σ_1_ is varied and σ_2_ is fixed at 2.8. The pink line is the proportion of runs where all three model parameters (κ_2_, λ_2_ and 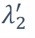) converged.

**Table S1.**
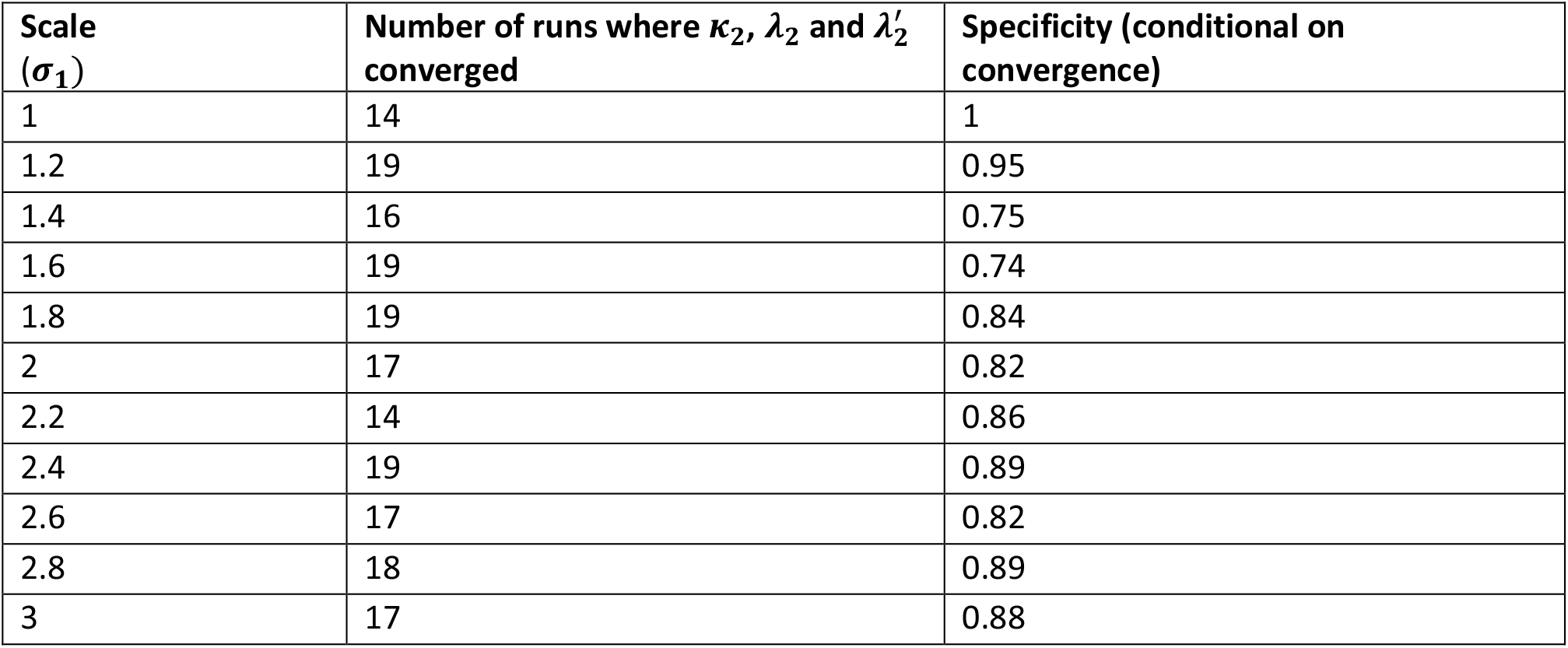
Specificity measured for each σ_1_ over 20 runs, after excluding runs where convergence was not achieved. Here, σ_2_ is fixed at 2.8.

| Scale ($\sigma_1$) | Number of runs where $\kappa_2$, $\lambda_2$ and $\lambda'_2$ converged | Specificity (conditional on convergence) |
| --- | --- | --- |
| 1 | 14 | 1 |
| 1.2 | 19 | 0.95 |
| 1.4 | 16 | 0.75 |
| 1.6 | 19 | 0.74 |
| 1.8 | 19 | 0.84 |
| 2 | 17 | 0.82 |
| 2.2 | 14 | 0.86 |
| 2.4 | 19 | 0.89 |
| 2.6 | 17 | 0.82 |
| 2.8 | 18 | 0.89 |
| 3 | 17 | 0.88 |

**Figure S4.**
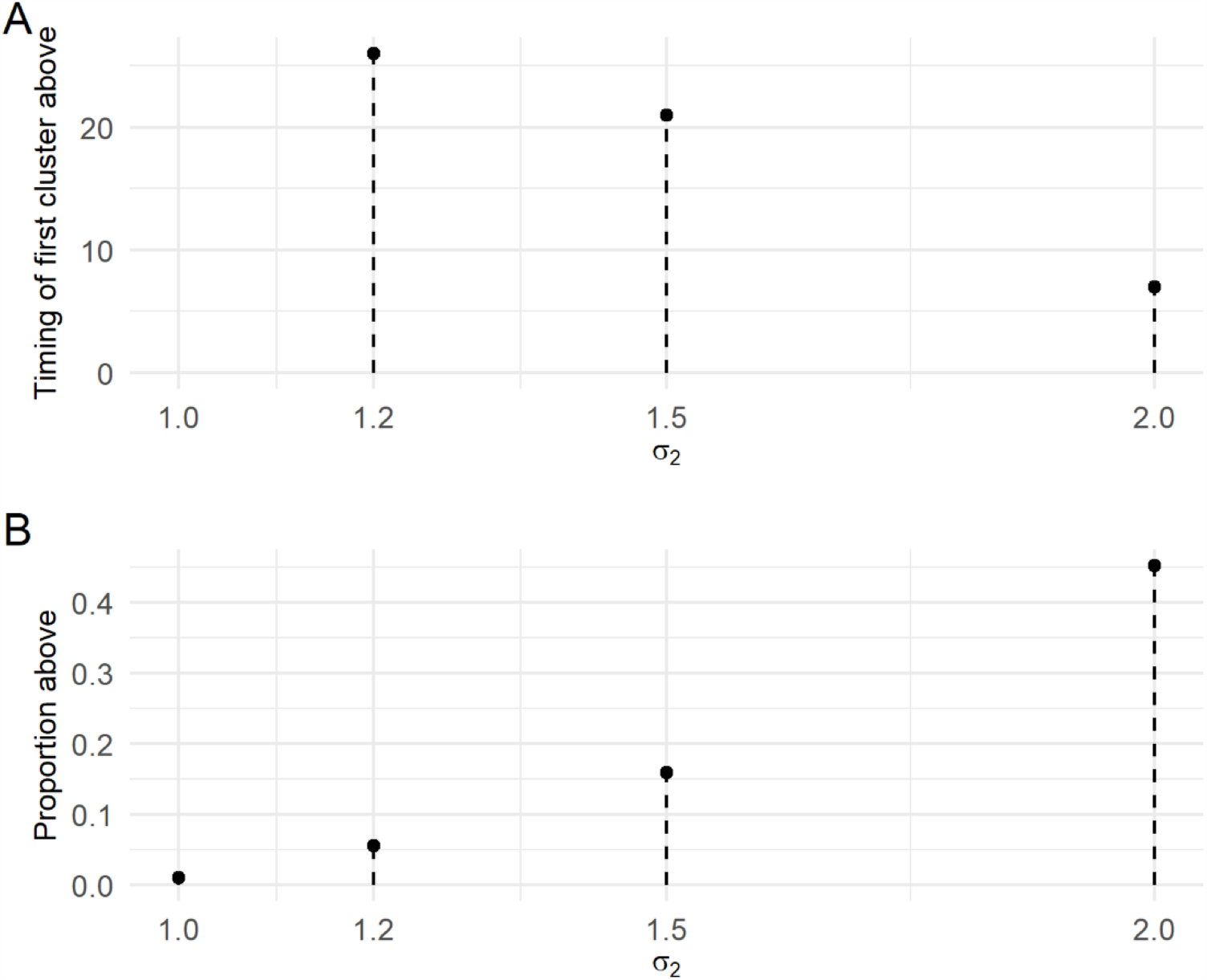
Results of the validation of the scenario where σ_1_ = 2.8 and σ_2_ varied from 1.2, 1.5 and 2. A shows the median of the timing of the first cluster of five consecutive observed reinfections, D_first_ above the projection interval for different values of σ_2_. B shows the proportion points above the projection interval for different values of σ_2_. Both metrics are for after the introduction of σ_2_ to the data. Runs that did not converge was excluded.

